# Evaluating the impact of a pulse oximetry remote monitoring programme on mortality and healthcare utilisation in patients with covid-19 assessed in Accident and Emergency departments in England: a retrospective matched cohort study

**DOI:** 10.1101/2021.11.25.21266848

**Authors:** T Beaney, J Clarke, A Alboksmaty, K Flott, A Fowler, JR Benger, P Aylin, S Elkin, A Darzi, AL Neves

## Abstract

**Objectives:** To identify the impact of a national pulse oximetry remote monitoring programme for covid-19 (COVID Oximetry @home; CO@h) on health service use and mortality in patients attending Accident and Emergency (A&E) departments.

**Design:** Retrospective matched cohort study of patients enrolled onto the CO@h pathway from A&E.

**Setting:** National Health Service (NHS) A&E departments in England.

**Participants:** All patients with a positive covid-19 test from 1^st^ October 2020 to 3^rd^ May 2021 who attended A&E from three days before to ten days after the date of the test. All patients who were admitted or died on the same or following day to the first A&E attendance within the time window were excluded.

**Interventions:** Participants enrolled onto CO@h were matched using demographic and clinical criteria to participants who were not enrolled.

**Main outcome measures:** Five outcome measures were examined within 28 days of first A&E attendance: i) death from any cause; ii) any subsequent A&E attendance; iii) any emergency hospital admission; iv) critical care admission; and v) length of stay.

**Results:** 15,621 participants were included in the primary analysis, of whom 639 were enrolled onto CO@h and 14,982 were controls. Odds of death were 52% lower in those enrolled (95% CI: 7%-75% lower) compared to those not enrolled on CO@h. Odds of any A&E attendance or admission were 37% (95% CI: 16-63%) and 59% (95% CI: 16-63%) higher, respectively, in those enrolled. Of those admitted, those enrolled had 53% (95% CI: 7%-76%) lower odds of critical care admission. There was no significant impact on length of stay.

**Conclusions:** These findings indicate that for patients assessed in A&E, pulse oximetry remote monitoring may be a clinically effective and safe model for early detection of hypoxia and escalation, leading to increased subsequent A&E attendance and admissions, and reduced critical care requirement and mortality.

## Background

The covid-19 pandemic has placed a huge demand on health systems around the world and led to an increase in use of digital technologies in public health responses and healthcare settings.[1–3] In the National Health Service (NHS) in England, embracing digital technologies was a priority even before the pandemic,[4] with system pressures from covid-19 driving an increased pace of adoption.[2] Remote monitoring devices have been highlighted as one of the technologies with the greatest potential impact on healthcare services,[4] with evidence suggesting that these can improve outcomes in selected patient groups.[5]

Early in the pandemic, it was recognised that hypoxia is a key prognostic marker and is strongly associated with mortality from covid-19.[6] A hallmark of covid-19 is the relative frequency of asymptomatic (‘silent’) hypoxia, making measurement of oxygen saturations a critical part of clinical assessment.[7,8] In England, NHS England and Improvement (NHSEI) launched the national COVID Oximetry @home (CO@h) programme in November 2020 to provide pulse oximeters to higher risk people diagnosed with covid-19 to support self-management and early recognition of hypoxia.[9] The intention of the programme, implemented in the community, was to accept referrals from primary care, NHS Test and Trace, ambulance services, and hospital emergency departments. In contrast, ‘COVID virtual wards’ operated from hospitals for those discharged following admission.[10]

Initial eligibility criteria for CO@h included adults aged 65 years or over, those designated as clinically extremely vulnerable (CEV), or where clinical judgment applied.[9] However, eligibility criteria varied across sites, and from February 2021, sites were encouraged to extend the age criteria to the 50+ age group. Those enrolled (“onboarded”) were encouraged to record three oximetry readings daily and advised to attend or call emergency services if the reading was 92% or less, or to contact primary care services for readings of 93-94%.[9] Implementation of the national programme built on an earlier pilot in four sites in England which has found it to be a safe pathway for people with covid-19.[11] However, evidence of the clinical effectiveness of the programme is lacking.

The aim of this study is to identify the effect of onboarding to the CO@h programme on 28-day mortality, subsequent A&E presentation, hospital admission, length of stay and critical care admission for patients assessed in A&E who do not require immediate hospital admission.

## Methods

This study used a retrospective matched cohort design. The eligible cohort included all people resident in England with a positive covid-19 test result between 1^st^ October 2020 and 3rd May 2021, who attended an NHS A&E department in England within a 14-day time window from 3 days before to 10 days after the date of their positive test. For all eligible patients, an A&E index date was created as the first A&E attendance date within the time window; for those onboarded, the attendance date on the same day or day prior to onboarding, within the time window was used. Any patient who was admitted or died on the same or following day to their index A&E attendance was excluded. Patients admitted to hospital in the 14 days before their index A&E attendance were also excluded, as were care home residents. Patients enrolled on the CO@h programme on the same or following day to their index A&E attendance (‘treated’) were matched to those not enrolled (‘controls’). Five outcomes were assessed, measured up to 28 days from index A&E attendance:

1. Death from any cause
2. One or more Accident and Emergency department (A&E) attendances
3. One or more emergency hospital admissions
4. One or more critical care admissions (of those admitted to hospital)
5. Total hospital length of stay in days, of those admitted who did not die within 28 days

### Data sources and processing

Data on patients onboarded to the CO@h programme was submitted from participating sites via NHS Digital’s Strategic Data Collection Service (SDCS).[12] Data on people with a positive covid-19 test was obtained from the Public Health England Second Generation Surveillance System,[13] which collates positive results from laboratories across England.[14] The date of a first positive covid-19 test was taken for each individual in cases where more than one test was recorded. A&E attendance data was provided through the Emergency Care Data Set.[15] Hospital admission data was provided from Hospital Episode Statistics (HES), linked to death registration data from the Office for National Statistics.[16] In patients admitted, total length of stay was capped at 28 days where a patient was discharged after the 28-day window. Patient demographics and chronic conditions were sourced from primary care data through the General Practice Extraction Service (GPES) Data for Pandemic Planning and Research (GDPPR).[17] Data were linked using a deidentified NHS patient ID.

Demographic data, including age, sex, ethnicity and lower layer super output area (LSOA) of residence were derived from GDPPR, or, if missing, from HES or ECDS. Deciles of the Index of Multiple Deprivation (IMD) 2019 were linked to LSOA of residence.[18] Data on clinically extremely vulnerable (CEV) status[19], body mass index (BMI), smoking and chronic conditions were derived from GDPPR. The following chronic conditions were included: hypertension, chronic cardiac disease, chronic kidney disease, chronic respiratory disease, dementia, diabetes, chronic neurological disease (including epilepsy), learning disability, malignancy/immunosuppression, severe mental illness, peripheral vascular disease and stroke/transient ischaemic attack (TIA). In each case, the latest codes were selected prior to the date of the positive covid-19 test, in order to exclude those potentially resulting from covid-19 infection. For the variables age, sex and ethnicity only, if no data were recorded prior to the date of the covid-19 test, the earliest data following the covid-19 test was used. In cases where the latest Systematised Nomenclature of Medicine Clinical Terms (SNOMED-CT) code indicated resolution of a condition, the condition was excluded. Further details of the datasets and processing are given in Appendix A, with the SNOMED-CT codes used in each condition cluster available in Appendix B.

### Statistical methods

Coarsened Exact Matching[20] was used, matching those onboarded with those not onboarded using the following variables: age category, sex, ethnicity, terciles of IMD score, BMI category, month of A&E index date, CEV status and days from covid-19 test to A&E index date (categorised as -3 to -1 days, 0 to 4 days, 5-10 days). The variables for inclusion in the model were chosen *a priori*. Patients with missing values for any of the matching covariates were excluded from analysis.

Logistic regression was used to estimate the odds ratio for each of the four binary outcomes in those onboarded compared to controls, applying stratum-specific weights from the matching algorithm to account for unequal stratum sizes. Negative binomial regression was used to estimate the treatment effect of the programme on length of stay. Analyses of length of stay excluded patients who died within the 28-day time window. Further details are given in the Appendix A.

Two sensitivity analyses were applied for each outcome to assess the robustness of inferences to the matching algorithm:

1. A doubly robust model, adjusted for all patient level covariates (the same covariates included in the matching, plus: smoking status and the twelve chronic diseases. In adjusted models, deciles of IMD score were used rather than the terciles used in matching.
2. A covariate-adjusted model, adjusted for the same variables as the doubly robust model, but without use of matching.

Analyses were conducted in the Big Data and Analytics Unit Secure Environment (BDAU SE), Imperial College. Python v3.9.5 and Pandas v1.2.3 were used in data manipulation. Matching was conducted in Stata v17.0, using the *cem* command.[20]

### Patient and public involvement

Patients or the public were not involved in the design, conduct or reporting of our research.

## Results

2,536,322 patients were identified with a positive covid-19 test between 1^st^ October 2020 and 3^rd^ May 2021. Of these, 220,473 (8.7%) attended A&E from 3 days before to 10 days after the positive test. After applying the exclusion criteria, 56,793 patients remained in the analysis, of whom 658 (1.2%) were onboarded to CO@h, and 56,135 (98.8%) were not onboarded (Figure 1).

**Fig 1:**
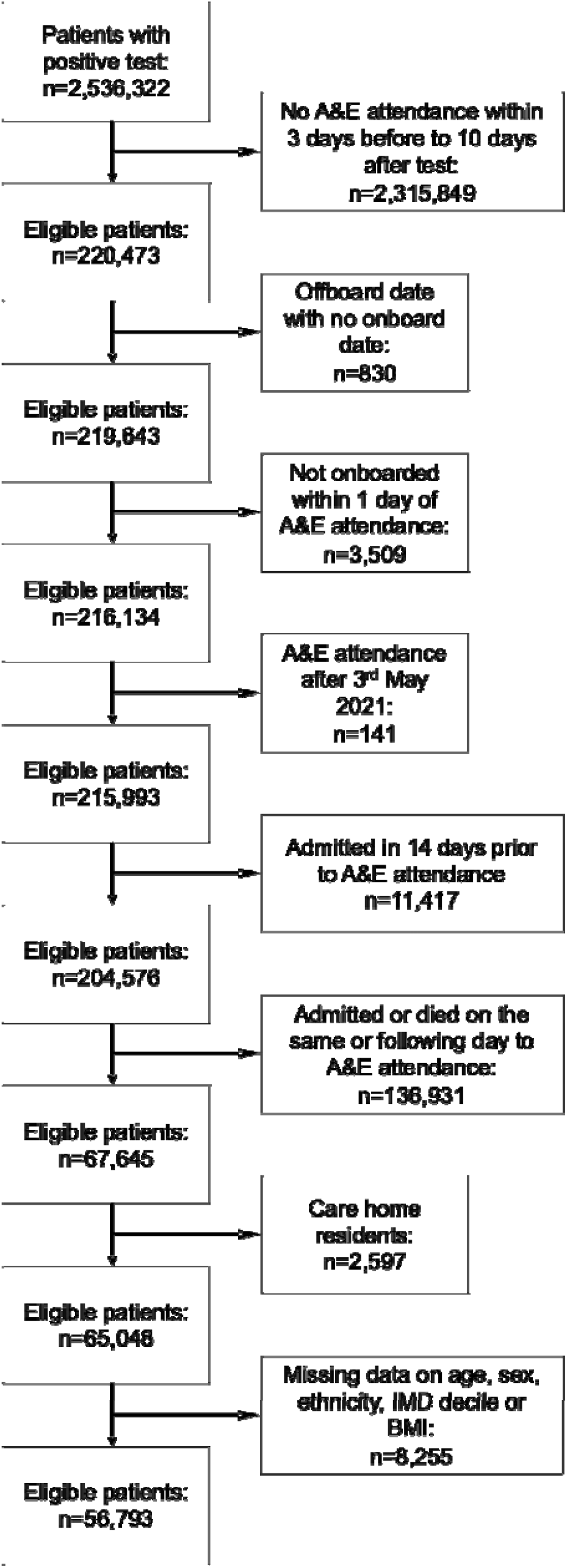
Flow chart for eligibility criteria for cohort.

639 (97.1%) onboarded patients were matched to 14,982 controls (representing 26.7% of total controls) giving a total of 15,621 in the primary analysis. The characteristics across each of the matching variables of the 19 unmatched and the 639 matched onboarded patients are given in Table 1. Those unmatched were more likely to be older, of non-white ethnic background, overweight and identified as CEV, although total numbers were small. There were no significant differences in the outcomes for matched compared to unmatched onboarded patients.

**Table 1:** Distribution of matching variables in unmatched and matched patients onboarded to CO@h after matching.

|  | Unmatched |  | Matched |  | p-value* |
| --- | --- | --- | --- | --- | --- |
|  | Number | Percentage | Number | Percentage |  |
| Age category (years) |  |  |  |  |  |
| 18-49 | 2 | 10.5% | 275 | 43.0% | 0.002 |
| 50-64 | 8 | 42.1% | 246 | 38.5% |  |
| 65-79 | 6 | 31.6% | 94 | 14.7% |  |
| 80 | 3 | 15.8% | 24 | 3.8% |  |
| Sex |  |  |  |  |  |
| Female | 11 | 57.9% | 331 | 51.8% | 0.600 |
| Male | 8 | 42.1% | 308 | 48.2% |  |
| Ethnicity |  |  |  |  |  |
| White | 5 | 26.3% | 495 | 77.5% | <0.001 |
| Asian/Asian British | 5 | 26.3% | 101 | 15.8% |  |
| Black/African/Caribbean/Black British | 1 | 5.3% | 17 | 2.7% |  |
| Mixed/Multiple ethnic groups | 4 | 21.1% | 7 | 1.1% |  |
| Other ethnic group | 4 | 21.1% | 19 | 3.0% |  |
| IMD tercile |  |  |  |  |  |
| 1 (most deprived) | 5 | 26.3% | 328 | 51.3% | 0.086 |
| 2 | 8 | 42.1% | 196 | 30.7% |  |
| 3 (least deprived) | 6 | 31.6% | 115 | 18.0% |  |
| Body Mass Index |  |  |  |  |  |
| Underweight | 1 | 5.3% | 1 | 0.2% | <0.001 |
| Healthy weight | 4 | 21.1% | 109 | 17.1% |  |
| Overweight | 10 | 52.6% | 216 | 33.8% |  |
| Obese | 4 | 21.1% | 313 | 49.0% |  |
| Clinically extremely vulnerable (CEV) status |  |  |  |  |  |
| Not CEV | 11 | 57.9% | 533 | 83.4% | 0.004 |
| CEV | 8 | 42.1% | 106 | 16.6% |  |
| Month of covid-19 test |  |  |  |  |  |
| September/October | 0 | 0.0% | 4 | 0.6% | 0.054 |
| November | 0 | 0.0% | 5 | 0.8% |  |
| December | 1 | 5.3% | 30 | 4.7% |  |
| January | 6 | 31.6% | 360 | 56.3% |  |
| February | 5 | 26.3% | 167 | 26.1% |  |
| March | 5 | 26.3% | 56 | 8.8% |  |
| April/May | 2 | 10.5% | 17 | 2.7% |  |
| Days from ED attendance to covid-19 test |  |  |  |  |  |
| -3 to -1 days | 2 | 10.5% | 27 | 4.2% | 0.403 |
| 0 to 4 days | 9 | 47.4% | 302 | 47.3% |  |
| 5 to 10 days | 8 | 42.1% | 310 | 48.5% |  |
| Outcome measures: |  |  |  |  |  |
| Deaths within 28 days | 1 | 5.3% | 9 | 1.4% | 0.176 |
| Patients with at least one ED attendance within 28 days | 3 | 15.8% | 192 | 30.0% | 0.18 |
| Patients with at least one admission within 28 days | 3 | 15.8% | 152 | 23.8% | 0.418 |
| Critical care use of those admitted | 0 | 0.0% | 9 | 1.4% | 0.664 |
| Total | 19 |  | 639 |  |  |

\*p-value from Chi-squared test

The odds of each binary outcome in those onboarded was compared to those not onboarded using logistic regression and negative binomial models were used for length of stay (Table 2). Patients onboarded had significantly lower odds of 28-day mortality (OR 0.48, 95% CI: 0.25-0.93; p=0.03) compared to those not onboarded. In contrast, those onboarded had a significant increase in the odds of any A&E attendance or hospital admission (OR 1.37, p<0.001 and OR 1.59, p<0.001, respectively). Among those admitted to hospital, patients previously onboarded had 0.47 times lower odds of receiving critical care (95% CI: 0.24-0.93, p=0.030). Of admitted patients, there was no significant difference in total length of stay from negative binomial regression models.

**Table 2:** Effect estimates associated with onboarding for each study outcome.

| Outcome | Odds ratio | Standard error | p-value | 95% confidence interval |  | Denominator |
| --- | --- | --- | --- | --- | --- | --- |
|  |  |  |  | Lower | Upper |  |
| 28-day mortality | 0.48 | 0.16 | 0.030 | 0.25 | 0.93 | 15,621 |
| Any A&E attendance within 28 days | 1.37 | 0.12 | <0.001 | 1.16 | 1.63 | 15,621 |
| Any hospital admission within 28 days | 1.59 | 0.15 | <0.001 | 1.32 | 1.91 | 15,621 |
| Any critical care use of those admitted | 0.47 | 0.16 | 0.030 | 0.24 | 0.93 | 2,272 |
|  | IRR* | Standard error | p-value | Lower | Upper | Denominator |
| Length of stay (LOS) of those admitted | 0.931 | 0.077 | 0.384 | 0.791 | 1.094 | 2,135 |

\*IRR = Incidence Rate Ratio

### Sensitivity analyses

A doubly robust model adjusted for all matching variables, in addition to the remaining patient level covariates that were not used in matching. The distributions of the remaining patient-level covariates were similar in the treated group and controls for the unmatched variables (Appendix A, Table A1). Effect sizes for the conditional odds ratios were slightly larger compared to those in the primary analysis, indicating a small degree of residual confounding in the unadjusted model, but these changes did not affect the inferences of the primary model (Table 3).

**Table 3:** Effect estimates associated with onboarding to CO@h for each study outcome, adjusted for patient level covariates.

| Outcome | Adjusted odds ratio | Standard error | p-value | 95% confidence interval |  | Denominator |
| --- | --- | --- | --- | --- | --- | --- |
|  |  |  |  | Lower | Upper |  |
| 28-day mortality | 0.43 | 0.16 | 0.025 | 0.20 | 0.90 | 15,327<sup>*</sup> |
| Any A&E attendance within 28 days | 1.41 | 0.13 | <0.001 | 1.18 | 1.69 | 15,621 |
| Any hospital admission within 28 days | 1.67 | 0.17 | <0.001 | 1.38 | 2.03 | 15,621 |
| Any level 2/3 care of those admitted | 0.43 | 0.15 | 0.019 | 0.21 | 0.87 | 2,249<sup>§</sup> |
|  | Adjusted IRR<sup>*</sup> | Standard error | p-value | Lower | Upper | Denominator |
| Length of stay (days) of those admitted | 0.989 | 0.078 | 0.884 | 0.85 | 1.15 | 2,135 |

\*IRR = Incidence Rate Ratio
&No deaths in November/April/May or mixed/multiple ethnic groups
§No critical care use of those admitted in April/May or mixed/multiple ethnic groups or underweight group

A second sensitivity analysis used a covariate adjusted model, without matching, to assess the robustness of inferences to the matching algorithm. This model used the full sample of 56,793 without applying matching weights and adjusted for all patient-level covariates (Table 4). Estimates of the conditional effect sizes were similar for all outcomes when compared to the primary model.

**Table 4:** Effect estimates associated with onboarding to CO@h for each study outcome from covariate adjusted models.

| Outcome | Adjusted odds ratio | Standard error | p-value | 95% confidence interval |  | Denominator |
| --- | --- | --- | --- | --- | --- | --- |
|  |  |  |  | Lower | Upper |  |
| 28-day mortality | 0.48 | 0.16 | 0.033 | 0.25 | 0.94 | 56,793 |
| Any A&E attendance within 28 days | 1.30 | 0.12 | 0.003 | 1.09 | 1.54 | 56,793 |
| Any hospital admission within 28 days | 1.57 | 0.15 | <0.001 | 1.30 | 1.91 | 56,793 |
| Any level 2/3 care of those admitted | 0.50 | 0.18 | 0.050 | 0.25 | 1.00 | 9,185 |
|  | Adjusted IRR<sup>*</sup> | Standard error | p-value | Lower | Upper | Denominator |
| Length of stay (days) of those admitted | 0.983 | 0.080 | 0.838 | 0.84 | 1.15 | 8,300 |

\*IRR = Incidence Rate Ratio

## Discussion

In a retrospective matched cohort study of patients with a positive covid-19 test assessed in A&E departments, those onboarded onto the CO@h programme had 52% lower odds of mortality within 28 days (OR: 0.48; 95% CI: 0.25-0.93, p=0.030) than those not onboarded. Those onboarded who were subsequently admitted also had lower odds of critical care use compared to those admitted who were not on the programme. In contrast, onboarding was associated with a significant increase in the odds of any subsequent A&E attendance (OR 1.37, p<0.001) or any emergency admission (OR 1.59, p<0.001) within 28 days.

Our findings support the aims of the CO@h programme which intends to enable early detection of hypoxia, and more timely clinical assessment and hospital admission, with resulting benefits for critical care admission and mortality. The magnitude of increase in emergency admissions was larger than that for subsequent A&E attendances, suggesting that presentations at A&E were clinically indicated and the programme did not create a burden of inappropriate demand. The results also highlight that the programme should not be viewed as a pathway to prevent hospital admission, but a pathway to support appropriate escalation and decision-making for assessment or admission.

There is limited pre-existing evidence for the effectiveness of pulse oximetry in health outcomes in patients with covid-19. A previous evaluation of the covid-19 pulse oximetry pilot programme in four sites in England found that none of those under 65 years and without long-term conditions died during the study, suggesting there were no safety concerns in lower risk patients, but without a control group to compare differences in clinical outcomes.[11] A recent systematic review identified thirteen studies of pulse oximetry monitoring in covid-19, but only two studies included control groups and only one of these compared health outcomes (Alboksmaty, A *et al*. Effectiveness and safety of pulse oximetry in remote home monitoring of COVID-19 patients: a systematic review. Unpublished, under review at The Lancet Digital Health). Gordon *et al* found lower odds of A&E presentation or readmission in patients discharged from hospital with remote monitoring, compared to those not enrolled.[21] However, the population discharged from hospital, analogous to the ‘virtual wards’ programme in England, is likely to be a very different patient group to those considered for community enrolment through the CO@h programme. A study of a telemonitoring service in Spain reported lower hospitalisations and mortality in those enrolled compared to the regional population, however, did not adjust for case-mix in the control population.[22]

In a second study of the CO@h programme by the same authors, analysing clinical outcomes at a population level, there was no effect on mortality and small increases in A&E attendances and hospital admissions following implementation of the CO@h programme.[23] This study found only 2.5% of eligible people nationally were enrolled, which although likely an underestimate, will dilute the effect of the programme at a population level. Taken together, these findings suggest that while the CO@h programme may promote timely detection of deterioration and escalation of care in those enrolled, the programme could not be provided at a wide enough scale to benefit the whole population as anticipated. Furthermore, none of the findings from either study indicate the programme causes harm, suggesting the programme is safe.

Further research is needed to identify other potential benefits and risks of the programme beyond clinical effectiveness and safety, including user experience of both patients and healthcare staff and the cost effectiveness of the programme. There is also a need to understand equity of the programme, both in terms of access to the programme and whether outcomes vary between different groups of people, which may allow for more effective targeting of the service.

### Strengths and limitations

A strength of this study is the use of data on those onboarded to the CO@h programme, as well as comprehensive data on all people resident in England with a positive covid-19 test, allowing matching of people onboarded to controls who would have been eligible for the programme but were not onboarded. Use of linked primary and secondary care data allowed the analysis to match using underlying patient risk factors, as well as month of test, to account for variation in outcomes over time[24] and days from test to A&E attendance, to account for differences in the course of disease over time. However, date of symptom onset was not available, so we were unable to account for confounding by time from symptoms to test. Matching criteria were determined *a priori* and use of different matching variables might impact on the findings. However, in two sensitivity analyses with different model specifications, inferences were the same as the primary model.

It is likely that there remains some residual confounding by disease severity. Decisions by clinicians in A&E on whether to admit or onboard patients will be influenced by disease severity at the time of presentation, and if there were systematic differences in severity in those onboarded compared to those not onboarded, the findings of our study will be biased. However, we believe there are two arguments that indicate it is unlikely that those onboarded had less severe disease than those not onboarded, and if anything, those onboarded are more likely to have had more severe disease. Firstly, if those onboarded had systematically lower severity, in the absence of any programme effect, we would expect admissions and mortality to reduce in parallel, rather than the divergent pattern seen in the results. Secondly, in the context of clinical assessment in A&E of whether to admit a patient, reassurance provided by a remote monitoring pathway may lower the threshold for A&E discharge, leading to the inclusion of a higher severity group in those onboarded. However, additional biases in patient selection may impact on the similarity of the two groups, such as through selection of those with greater digital literacy or exclusion of those who were already monitoring their oxygen saturations using personally purchased oximeters.

The findings of this study apply to the subset of people with covid-19 who were reviewed in A&E but did not require immediate admission, and who did not die within one day of A&E attendance. As a result, the findings may not be generalisable to the wider population eligible for the programme, for example, those presenting to primary care, or at an earlier or later stage of disease. The outcome estimates also exclude those with the most severe disease, who were admitted or died within 24 hours of A&E attendance and so may not be comparable to estimates from other studies.

The CO@h programme is not a homogenous programme, with variation in the type of model implemented, and it is unlikely that a single effect estimate will be representative across all sites.[25] Clinical decision-making with regards to onboarding will also be specific to both A&E department and CO@h site, particularly given the emphasis on clinical judgment to determine eligibility.[9] Given the small number of onboarded patients in our study, data was insufficient to match on A&E department, or to examine outcome measures within single sites. There may also be groups of patients for whom the programme is more suitable, and concern that pulse oximetry is more likely to be falsely reassuring in people with black or brown skin,[8] however, due to the small sample size, subgroup analyses were outside the scope of the study.

## Conclusion

The CO@h programme, implemented in England from November 2020, sought to enable early recognition and intervention for people with covid-19-induced hypoxia. The results of this study support the aims of the programme in the subset of patients assessed in A&E and found that those enrolled had significantly lower odds of mortality and requirement for critical care admission within 28 days, and higher odds of subsequent A&E attendance and emergency hospital admission compared to those not enrolled, suggesting early recognition of hypoxia and appropriate escalation of care. These findings indicate that for individual patients, pulse oximetry remote monitoring may be clinically effective in reducing mortality and the need for critical care admission.

## Data Availability

The patient level data used in this study are not publicly available but are available to applicants meeting certain criteria through application of a Data Access Request Service (DARS) and approval from the Independent Group Advising on the Release of Data.

## Acknowledgements

The authors would like to thank Hutan Ashrafian, Gianluca Fontana, Saira Ghafur, Melanie Leis and Mahsa Mazidi for their input and support. Data management was provided by the Big Data and Analytical Unit (BDAU) at the Institute of Global Health Innovation (IGHI), Imperial College London. This work was funded by NHS England and supported by the National Institute for Health Research (NIHR) Imperial Patient Safety Translation Research Centre. Infrastructure support was provided by the NIHR Imperial Biomedical Research Centre (BRC). JC acknowledges support from the Wellcome Trust (215938/Z/19/Z). The study funder(s) did not play a role in study design; in the collection, analysis, and interpretation of data; in the writing of the report; and in the decision to submit the article for publication. In addition, researchers were independent from funders, and all authors had full access to all of the data included in this study and can take responsibility for the integrity of the data and the accuracy of the data analysis.

## Ethics approval

The work was conducted as a national service evaluation of the CO@h programme, approved by Imperial College Health Trust on 3^rd^ December 2020. Data access was approved by the Independent Group Advising on the Release of Data (IGARD; DARS-NIC-421524-R0Y3P) on 15^th^ April 2021.

