## Supplementary appendix A for "Evaluating the impact of a pulse oximetry remote monitoring programme on mortality and healthcare utilisation in patients with covid-19 assessed in Accident and Emergency departments in England: a retrospective matched cohort study"

**Data cleaning**

Note: the described datasets were used for several distinct analyses by the study team, with data cleaning rules the same or similar between studies. For this reason, some of the text in the appendices may be identical to that of other published articles by the study authors on the same data source.

*Covid-19 testing data*

Testing data was provided through the Public Health England Second Generation Surveillance System (SGSS). This dataset captures routine laboratory data on infectious diseases for England, including Covid-19, with all diagnostic laboratories required to notify positive test results within 24 hours.^1^ Data included 3,251,225 tests performed from 1^st^ October 2020 to 30^th^ June 2021, inclusive.

Provided data included test date and result date. 99% of results were reported within 5 days of the test, and 6,544 (0.2%) were reported more than 7 days from date of test. Where the result data occurred before the test date. In 1,603 cases, the result date was recorded as prior to the test date. In these instances, where the difference between the testing date and reporting date was 7 days or less, the test date and reporting date were swapped. In the 191 instances where reporting date was more than 7 days before the testing date, the test was excluded.

For this analysis, only tests performed up to 3^rd^ May 2021 were included (after swapping test and result dates where applicable), given that secondary care data was available up until the end of May 2021. Of the 2,928,802 positive Covid-19 tests, 2,352,390 (80.3%) were from Pillar 2 testing, 561,852 (19.2%) from Pillar 1, and 14,560 (0.5%) from Pillar 4.^2^ Test type was recorded for Pillar 2 tests only, and of these, 2,250,288 (95.7%) were Polymerase Chain Reaction (PCR) tests, 102,102 (4.3%) were lateral flow tests.

For this analysis, only the first positive test was taken where more than one was recorded for any given individual, resulting in a total eligible population of 2,536,322.

*COVID Oximetry @home (CO@h) programme data*

Data on patients enrolled (‘onboarded’) onto the CO@h programme were submitted directly from CO@h sites via NHS Digital’s Strategic Data Collection Service.^3^ Data included a deidentified NHS patient ID of the patient onboarded, along with the date of onboarding to and offboarding from the programme. Any patient with an offboarding date but no onboarding date were excluded from the analyses.

*Primary care data*

Primary care data came from the General Practice Extraction Service (GPES) Data for Pandemic Planning and Research (GDPPR).^4^ Data included month and year of birth, sex, ethnicity, Lower Layer Super Output Area (LSOA) of residence, a marker for Clinically Extremely Vulnerable (CEV) status, and a marker for residence in a care home. LSOA was used to link to 2019 deciles of Index of Multiple Deprivation (IMD).^5^ Age was calculated from date of positive Covid-19 test, assuming a birthdate on the 15^th^ day of the month.

Entries include a date to which each journal item applies, and a date on which the journal item was recorded. The former was used in priority, but where missing, was replaced with the journal item recording date. For LSOA, CEV status and care home residence, only entries occurring up to the date of positive Covid-19 test were included. For month and year of birth, sex and ethnicity, if no entry were included prior to the date of Covid-19 test, then the earliest recorded entry after the test was included.

*Secondary care data*

Data on hospital admissions came from the Hospital Episode Statistics (HES) data set up to 31st May 2021, linked to Office for National Statistics (ONS) data on death registrations up to 5th July 2021.^6^ Entries were excluded where missing admission dates, provider Trust code, or patient deidentified ID.

Where multiple admission episodes were recorded within a spell, a single spell start and end date were created. Non-emergency hospital admissions were excluded from analyses. A binary indicator was created for any admission within 28 days of positive Covid-19 test. A second indicator was created for death (of any cause) within 28 days of positive Covid-19 test. Critical care admissions were defined as an admission episode for any level 2 or level 3 care within the defined spell.

Data on accident and emergency (A&E) attendances came from the Emergency Care Data Set (ECDS).^7^ Only A&E presentations to NHS providers were selected (Trust codes beginning with ‘R’). The A&E attendance date, departure date and admission date (if subsequently admitted) are included. In cases where attendance date was recorded as being after the departure date, the attendance date was set to the departure date, if departure date was equal to the admission date. Otherwise, attendance date was assumed to be correct.

Where multiple A&E attendances were recorded on the same day, a single attendance was kept for each patient, prioritising in turn:

1. Any attendance associated with an admission
2. Earliest time of attendance
3. Earliest time of departure

A binary indicator was created for one or more A&E attendances within 28 days of a positive Covid-19 test.

Where age was missing from GDPPR, it was derived from month and year of birth in HES, or if also missing in HES, derived from month and year of birth in ECDS, using the same approach as for GDPPR. Where LSOA was missing from GDPPR, it was derived from HES/ECDS. CCG was derived first from testing data, and if missing, from CO@h programme data, followed by GDPPR/HES/ECDS if missing.

*Co-morbidities*

SNOMED codes were included in the GDPPR dataset pertaining to specific SNOMED code cluster reference sets provided by NHS Digital.^4^ 6,485 unique codes were identified from GDPPR. Codes were reviewed manually by authors TB and JC and removed if not relevant or assigned to the minimal number of relevant code clusters.

SNOMED reference clusters were aggregated into hierarchies of similar conditions. Codes in each higher-order cluster were then reviewed to ensure groupings of relevant codes and twelve relevant chronic disease categories were selected: hypertension, chronic cardiac disease, chronic kidney disease, chronic respiratory disease, dementia, diabetes, chronic neurological disease (including epilepsy), learning disability, malignancy/immunosuppression, severe mental illness, peripheral vascular disease and stroke/transient ischaemic attack (TIA). Categories for chronic respiratory disease, diabetes, epilepsy, malignancy/immunosuppression and severe mental illness included relevant medication codes. Broad diagnostic categories of diagnoses were chosen, as certain medications were not diagnostic of more granular diagnostic categories (for example, use of a long-acting bronchodilator/inhaled corticosteroid in both COPD and asthma) A full list of codes within each diagnostic category are available in Appendix B.

For each patient in GDPPR, all relevant diagnostic codes prior to the study index date (date of positive Covid-19 test) were considered diagnostic. In cases where the latest SNOMED code indicated resolution of a condition (e.g. ‘Atrial fibrillation resolved (finding)’), then the diagnosis was excluded for that patient. SNOMED codes relating to drug codes were only included up to 2 years prior to the index date.

*BMI categorisation*

SNOMED codes for BMI were either diagnostic categories (eg ‘Body mass index 30+ - obesity (finding)’ or value codes (e.g. ‘Body mass index (observable entity)’). Values were extracted and BMI was categorised according to the standard World Health Organisation classification of underweight (<18.5 kg/m^2^), healthy weight (18.5-24.9 kg/m^2^), overweight (25.0-29.9 kg/m^2^) and obese (≥30.0 kg/m^2^). Value codes outside of the range 5.0-100.0 kg/m2 were excluded. SNOMED codes which spanned more than one category (e.g. ‘Increased body mass index (finding)’) and child BMI categories were also excluded.

*Smoking categorisation*

Smoking status was categorised into ‘never-smoker’, ‘ex-smoker’ and ‘current smoker’ according to the latest SNOMED code prior to and including the index date. For any patient where the latest SNOMED code indicated ‘never-smoker’, but a prior record indicated active smoking, then the patient was re-categorised as ‘ex-smoker.

**Statistical analysis**

Coarsened Exact Matching was used to match onboarded patients (‘treated’) and controls, using the *cem* command in Stata following the approach defined by Blackwell et al (2010).^8^ Patients with any of the matching variables missing (8,255; 12.7%) were dropped for the analysis, resulting in a total of 56,135 controls and 658 treated.

The following variables were chosen *a priori* for inclusion in the matching algorithm:

- Age category (<50, 50-64, 65-79, 80+ years)
- Sex (female or male)
- Ethnicity (using the five ONS categories: White, Asian/Asian British, Black/African/Caribbean/Black British, Mixed/Multiple ethnic groups, Other ethnic groups)
- Terciles of IMD score
- BMI category (underweight, healthy weight, overweight, obese)
- Month of ED index date (September-October 2020 & April-May 2021 were combined due to small numbers in September and May)
- Clinically Extremely Vulnerable (CEV) status (yes/no)
- Days from Covid test to A&E index date (cuts applied at: -3 to -1; 0 to 4; 5 to 10 days)

Logistic regression was used to estimate the treatment effect of the programme separately for each of the four binary outcomes occurring in the time period from A&E index date to 28 days:

- Death from any cause
- Any ED attendance (excluding the ED index attendance)
- Any emergency hospital admission
- Any critical care (level 2 or level 3) admission, of those admitted

Negative binomial regression was used to estimate the treatment effect of the programme on length of stay in days, for those admitted within 28 days. Total length of stay was capped at 28 days where a patient was discharged after the 28-day window, and analyses of length of stay excluded patients who died within the 28-day time window. The overdispersion parameter was calculated and the distribution of length of stay was found to be overdispersed compared to a Poisson process in all models, indicating superior model fit for the negative binomial model compared to the Poisson model. Stratum-specific weights from the matching algorithm were applied to all regression models to account for an unequal ratio of controls to onboarded patients across each stratum.

Two sensitivity analyses were applied to the models for each of the logistic and negative binomial regression models:

1. A doubly robust model, adjusted for all patient level covariates (matching variables, plus: smoking status, hypertension, chronic cardiac disease, chronic kidney disease, chronic respiratory disease, dementia, diabetes, chronic neurological disease (including epilepsy), learning disability, malignancy/immunosuppression, severe mental illness, peripheral vascular disease and stroke/TIA).
2. A covariate-adjusted model, adjusted for the same variables as the doubly robust model, but without use of the matching.

In adjusted models, IMD score was treated as deciles to give greater granularity from the terciles used in matching.

**Table A1: Proportions of patient-level covariates not used in matching, between onboarded patients and controls**

| **Covariate** | **Percentage (%)** | |
| --- | --- | --- |
|  | **Controls** | **Onboarded** |
| **Smoking status** | | |
| Never smoker | 56.4 | 56.7 |
| Ex-smoker | 27.9 | 28.3 |
| Current smoker | 15.8 | 15.0 |
| **Co-morbidities** | | |
| Hypertension | 24.6 | 25.8 |
| Chronic cardiac disease | 10.5 | 9.7 |
| Chronic kidney disease | 1.0 | 1.1 |
| Chronic respiratory disease | 30.7 | 37.1 |
| Dementia | 0.7 | 0.3 |
| Diabetes | 17.5 | 18.3 |
| Chronic neurological disease (including epilepsy) | 4.7 | 5.2 |
| Learning disability | 0.7 | 0.8 |
| Malignancy or immunosuppression | 10.1 | 10.6 |
| Severe mental illness | 3.9 | 2.8 |
| Peripheral vascular disease | 1.2 | 1.6 |
| Stroke or TIA | 3.5 | 3.3 |

6.	Digital, N. Hospital Episode Statistics (HES). https://digital.nhs.uk/data-and-information/data-tools-and-services/data-services/hospital-episode-statistics.

7.	NHS Digital. Emergency Care Data Set (ECDS). https://digital.nhs.uk/data-and-information/data-collections-and-data-sets/data-sets/emergency-care-data-set-ecds.

8.	Blackwell, M., Iacus, S., King, G. & Porro, G. Cem: Coarsened Exact Matching in Stata. *Stata J.* **9**, 524–546 (2009).
